# An electrocardiogram-based machine learning model for distinguishing complete Kawasaki disease

**DOI:** 10.64898/2026.04.30.26352183

**Authors:** Takashi Nakano, Kazuyoshi Saito, Kasumi Noda, Yumiko Asai, Arisa Kojima, Hidetoshi Uchida, Yoshimi Ohira, Hiroyasu Ito, Jun-ichi Kawada, Tetsushi Yoshikawa

## Abstract

Kawasaki disease (KD) is a systemic vasculitis in young children, and early diagnosis remains challenging when clinical features are incomplete or overlap with those of other febrile illnesses. Because electrocardiography (ECG) is noninvasive and widely available, we investigated whether ECG-derived features could help distinguish complete KD from pediatric patients with fevers. We conducted a single-center retrospective study of hospitalized febrile children aged 1–8 years who underwent digital 12-lead ECG recording during the initial evaluation. Five amplitude features and six timing features extracted from the ECG were used to develop a logistic regression model to distinguish between complete KD and other febrile illnesses. The model discriminated between the KD and non-KD groups in the validation dataset. The prediction score was not significantly correlated with the age and body temperature. S-wave amplitude, the RR interval, and P-and Q-wave amplitudes were suggested to contribute to discrimination. These findings suggest that ECG-derived features may provide adjunctive information for distinguishing complete KD from other febrile illnesses.

**Author Summary:** Kawasaki disease is an inflammatory illness in young children that can lead to coronary artery complications if treatment is delayed. Early diagnosis is often difficult because its initial symptoms overlap with those of many common febrile illnesses. We investigated whether a routine 12-lead electrocardiogram (ECG), which is noninvasive, rapid, and widely available, contains information that can help distinguish complete Kawasaki disease from other febrile conditions. We retrospectively analyzed digital ECGs from hospitalized febrile children and extracted waveform amplitude and timing features. Using these features, we built a logistic regression model and evaluated it in a temporally separate validation cohort. The model distinguished patients with Kawasaki disease from patients with fever. P-, Q-, and S-wave amplitudes and the RR interval were repeatedly selected as important contributors, suggesting that both waveform morphology and heart-rate-related information may be relevant. These findings indicate that ECG-derived features may provide useful adjunctive information during the clinical assessment of complete Kawasaki disease.

## Introduction

Kawasaki disease (KD) is a systemic vasculitis that primarily affects infants and young children. If appropriate treatment is delayed, serious cardiovascular complications, including coronary artery aneurysm formation, may occur. Clinical diagnosis is based on fever and a constellation of principal clinical features [1,2]. However, atypical or evolving presentations are common in the early phase of illness, and KD can be difficult to distinguish from other pediatric febrile illnesses [3,4]. Establishing objective, noninvasive adjunctive diagnostic markers therefore remains an important challenge in the clinical management of KD.

Electrocardiography (ECG) is a simple, noninvasive examination that can be repeated easily and is widely used in pediatric practice. ECG changes in KD have traditionally been considered nonspecific, but studies focusing on abnormalities in ventricular repolarization have accumulated in recent years. Several studies have reported that variation in ventricular repolarization may reflect inflammatory burden, suggesting an association between systemic inflammation and ECG findings in KD [5–8]. However, most previous studies focused on specific indices, and attempts to make comprehensive use of information from the entire ECG have been limited.

Recent advances in machine learning have expanded the use of data-driven methods for disease classification. High diagnostic accuracy has been reported in adults for arrhythmias and ischemic heart disease [9–11]. Most machine learning studies on KD have focused on blood test results or inflammation-related biomarkers [12–15]. A recent systematic review of machine learning for KD also found that the included diagnostic models were based primarily on common clinical data [12]. ECG-based studies in KD remain limited.

In this study, we investigated whether ECG features extracted from 12-lead ECG data could be used to distinguish complete KD from other pediatric febrile diseases using machine learning. Because ECGs are digitally native, widely available, noninvasive, rapidly acquired, and relatively inexpensive, an ECG-based model could be clinically useful as an adjunctive triage aid if it provides incremental information beyond routine assessment.

## Materials and Methods

### Study design and population

This retrospective observational study was designed with temporal validation. We included febrile (≥37.5°C) patients aged 1–8 years who were hospitalized at Fujita Health University Hospital between 2014 and 2024 and had digital 12-lead ECG data recorded. The Kawasaki disease (KD) group was limited to cases diagnosed as complete KD after review of the medical records by at least three pediatricians. Complete KD was defined as fulfillment of at least five of the six principal diagnostic criteria for KD, according to the sixth revised edition of the diagnostic guidelines. The control group (non-KD) consisted of patients with a fever but not KD, including infections (n = 38), perioperative conditions (n = 29), neurologic disorders (n = 14), and other conditions.

The discovery dataset used for model development and lead exploration comprised patients hospitalized from 2014 to 2021, whereas the validation dataset comprised patients hospitalized from 2022 to 2024 (Table 1).

**Table 1.**
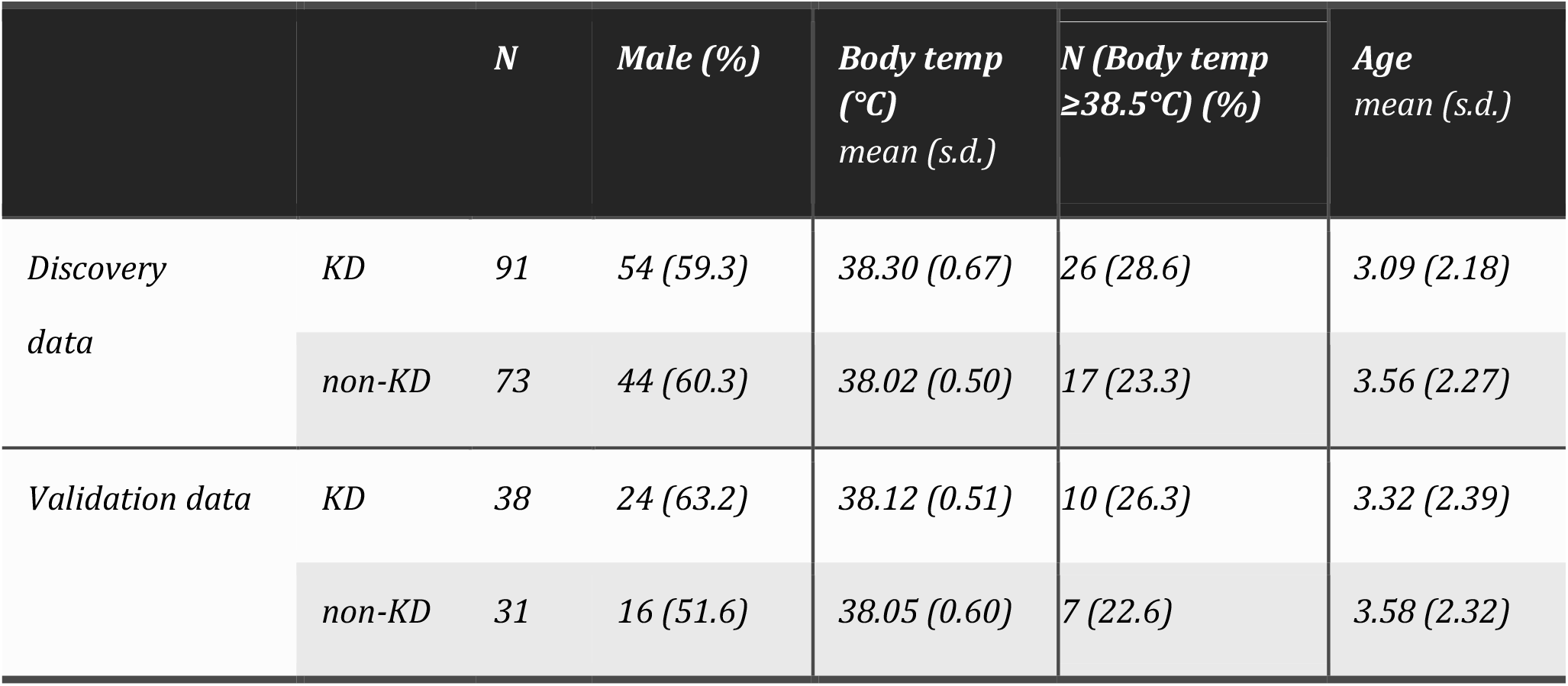
Cohort characteristics of the discovery and validation datasets.

This study was approved by the institutional ethics committee (approval number: HM25-281). Because of the retrospective study design, informed consent was obtained through an opt-out approach.

### ECG acquisition and preprocessing

12-lead ECGs recorded using electrocardiographs (FDX-4520, FCP-7541, VS-3000TE, FCP-8700, FCP-8800, and FCP-9800; Fukuda Denshi Co., Ltd., Tokyo, Japan) under standard conditions for 10 seconds with a sampling frequency of 500 Hz were used. For KD cases, ECGs recorded during the acute phase, before treatment initiation were analyzed. ECGs from the control group were obtained at the time of the initial evaluation. After anonymization, ECG data underwent preprocessing, including noise reduction with a band-pass filter (1–40 Hz) and a 60-Hz notch filter, followed by baseline correction.

### Feature extraction

Fiducial point estimation and feature extraction from ECG waveforms were performed using ECGdeli, an open-source ECG delineation toolbox for MATLAB [16]. As amplitude features, the peak amplitudes of the P, Q, R, S, and T waves were calculated. Candidate leads comprised the eight leads I, II, and V1–V6. The III lead and augmented limb leads (aVR, aVL, and aVF) were excluded because they can be derived as linear combinations of other leads. Timing features included the PR interval, QRS duration, RR interval, QT interval, QT dispersion (QTd; maximum QT minus minimum QT across the 12 leads), and T_peak_-T_end_ dispersion (Tped).

### Model development

Model development was performed in MATLAB R2025b. A logistic regression model was used for binary classification of KD versus non-KD. To reduce both class imbalance and age-related confounding, random downsampling was performed within prespecified age strata so that the KD and non-KD groups contained equal numbers of patients at each integer age. This random downsampling procedure was repeated 20 times, and performance metrics are reported as mean ± standard deviation. Model development and evaluation were conducted in two stages: (1) exploratory 5-fold cross-validation (CV) within the discovery dataset and (2) temporal validation in which a model trained on the downsampled discovery dataset was applied to the validation dataset. For each stage, all features were standardized by z-score using the training data, and the test data were transformed using the same parameters. Patients were classified as KD when the predicted probability from the logistic regression model exceeded 0.5. Performance metrics were accuracy, sensitivity, specificity, and the area under the receiver operating characteristic curve (AUC).

### Exhaustive feature exploration

To explore which leads yielded the highest classification performance among the amplitude features, we evaluated all possible combinations in which one lead was selected from the eight candidate leads for each of the P, Q, R, S, and T waves (8^5^ = 32,768 combinations). For each combination, a logistic regression model built using the amplitude features and the timing features (PR interval, QRS duration, RR interval, QT interval, QTd, and Tped) was evaluated by 5-fold CV within the discovery dataset as exploratory lead screening. In addition, for the total set of 11 features consisting of the five amplitude features derived from the representative leads and the six timing features, we also exhaustively evaluated all 2^11^-1 = 2,047 feature subsets to explore which combinations were repeatedly associated with higher prediction performance.

## Results

### Lead selection and classification performance

We first explored which of the eight candidate leads was most informative for the amplitude of each wave (P, Q, R, S, and T) in distinguishing complete KD from non-KD. We enumerated all 32,768 possible combinations obtained by selecting one lead for each wave and calculated the AUC of a logistic regression model for each combination (Figure 1A). The highest AUC values reached approximately 0.8, and the top-ranked combinations by AUC also showed relatively high accuracy (Figure 1B).

**Figure 1.**
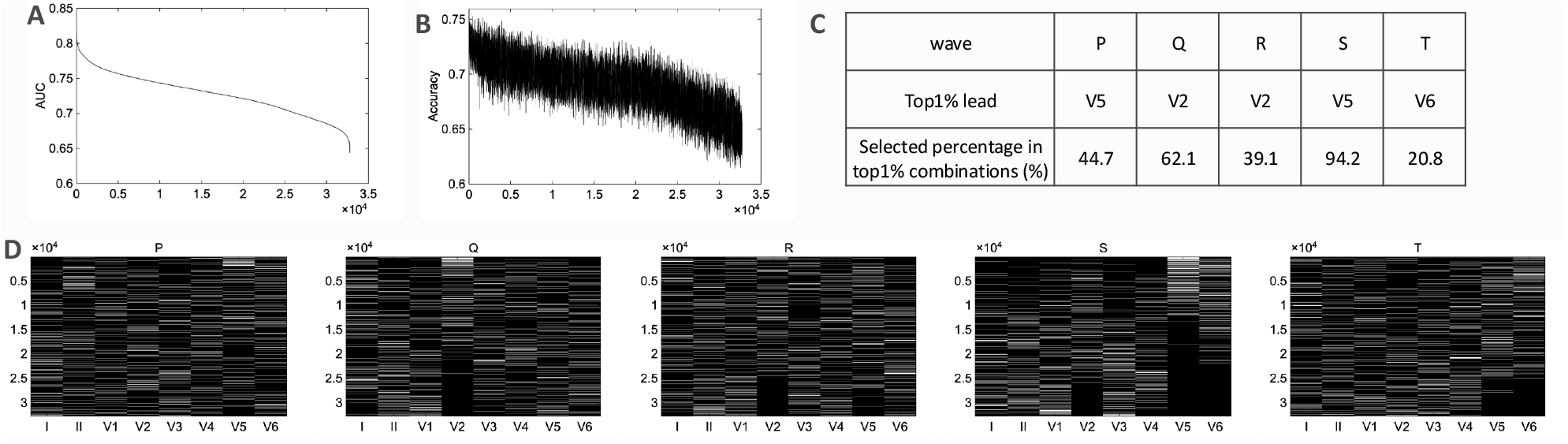
Lead exploration for amplitude features. (A) AUC values for all combinations (8^5^ = 32,768). (B) Distribution of accuracy across all combinations, ordered by AUC rank. (C) The most frequently selected lead and its selection frequency among the top 1% of combinations ranked by AUC. (D) Heatmap showing the distribution of selected leads for each wave (P, Q, R, S, and T) across AUC-ranked combinations.

When the frequency of lead occurrence was tabulated among the top 1% of combinations ranked by AUC, the representative leads most frequently selected for each wave were V5 for the P wave, V2 for the Q wave, V2 for the R wave, V5 for the S wave, and V6 for the T wave (Figure 1C). In particular, V5 was selected for the S wave in 94.2% of combinations, V2 was selected for the Q wave in 62.1%, and V5 was selected for the P wave in 44.7% indicating relatively stable selection patterns. In contrast, the proportions of the most frequent leads were lower for the R wave (39.1%) and T wave (20.8%), indicating less concentration on a single lead (Figure 1D).

Using the representative leads identified in Figure 1 (P: V5, Q: V2, R: V2, S: V5, and T: V6), we constructed five amplitude features and combined them with the timing features (QTd, PR interval, QT interval, Tped, QRS duration, and RR interval), yielding a total of 11 features for logistic regression-based binary classification of KD versus non-KD. Models trained on 20 age-matched downsampled versions of the discovery dataset were applied to the validation dataset. As a result, the mean performance was an accuracy of 0.703 ± 0.017, sensitivity of 0.684 ± 0.023, specificity of 0.726 ± 0.022, and AUC of 0.737 ± 0.015 (Figure 2).

**Figure 2.**
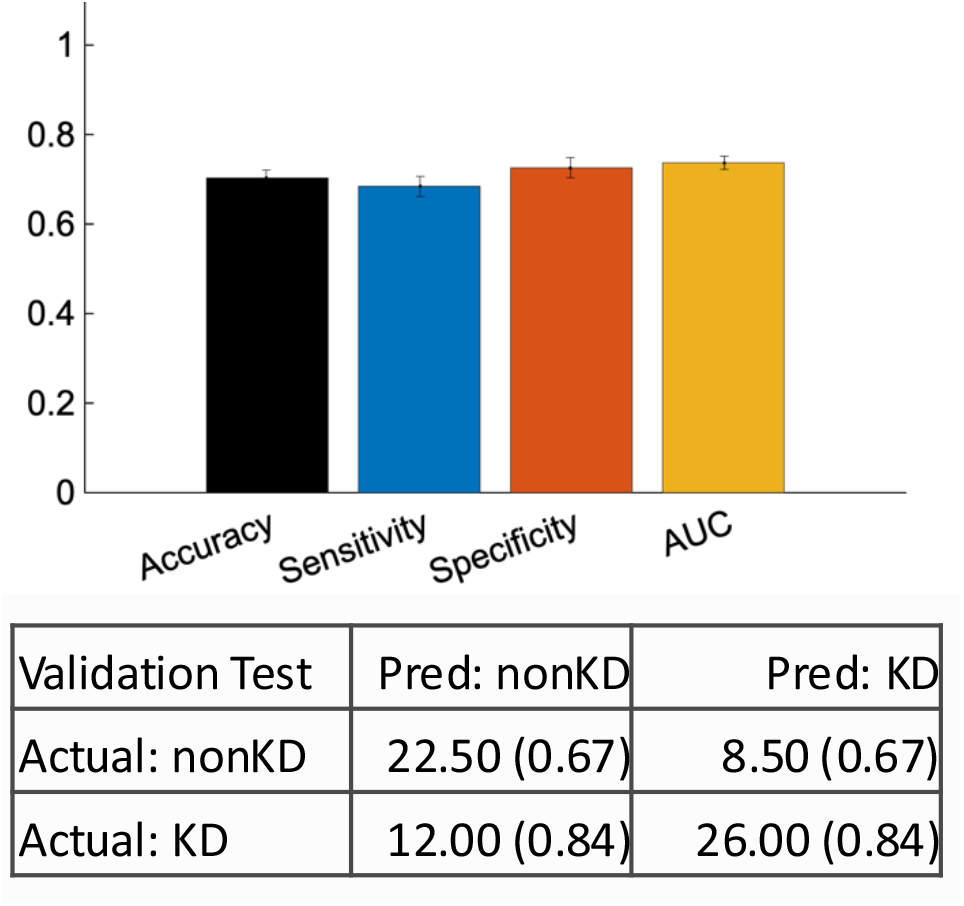
Classification performance of the 11-feature logistic regression model. Performance metrics include accuracy, sensitivity, specificity, and AUC. The confusion matrix shows mean counts across 20 age-matched downsampling iterations, with standard deviations.

We then investigated which features contributed to classification. Figure 3 shows the logistic regression coefficients of the model trained on the 20 age-matched downsampled discovery datasets. Among the coefficients, the RR interval showed the largest negative value, and P-, Q-, S- and T-wave amplitudes, and QT interval also showed a relatively large contribution (Figure 3A). In addition, in the exhaustive evaluation of all 11-feature subsets, S-wave amplitude and the RR interval were included in 100% of the top 1% AUC-ranked combinations, followed by P-wave amplitude and Q-wave amplitude, each of which was included in 90% and 85% of these combinations, respectively. T-wave amplitude, R-wave amplitude, PR interval, QRS duration, and the remaining timing features were included less frequently (Figure 3B). These results indicated the amplitudes of P, Q, and S waves and the RR interval contribute to discrimination between KD and non-KD.

**Figure 3.**
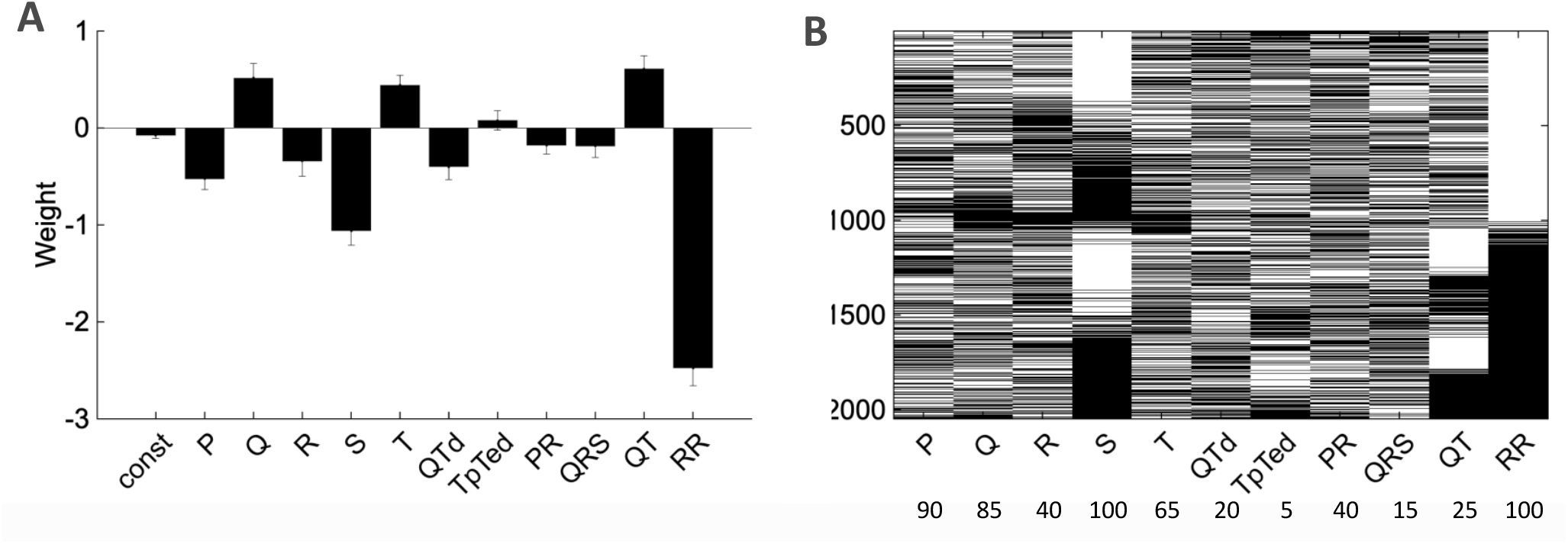
Contribution of the 11 features. (A) Mean logistic regression coefficients across 20 age-matched downsampled discovery datasets. Error bars indicate standard deviations. (B) Results of the exhaustive search of feature combinations. White indicates selected features, and the numbers at the bottom indicate the proportion of top 1% AUC-ranked combinations that included each feature.

Because KD is a pediatric disease and age can substantially affect ECG findings, we evaluated the potential confounding effect of age (Figure 4). In the discovery dataset, performance metrics varied across age groups, but an AUC greater than 0.6 was maintained in many age groups (Figure 4A). Similar variation across age was observed in the validation dataset (Figure 4B). The prediction score was not significantly correlated with age (r = −0.14, p = 0.25) (Figure 4C), suggesting that the discriminative information captured by the model was not explained solely by differences in age distribution.

**Figure 4.**
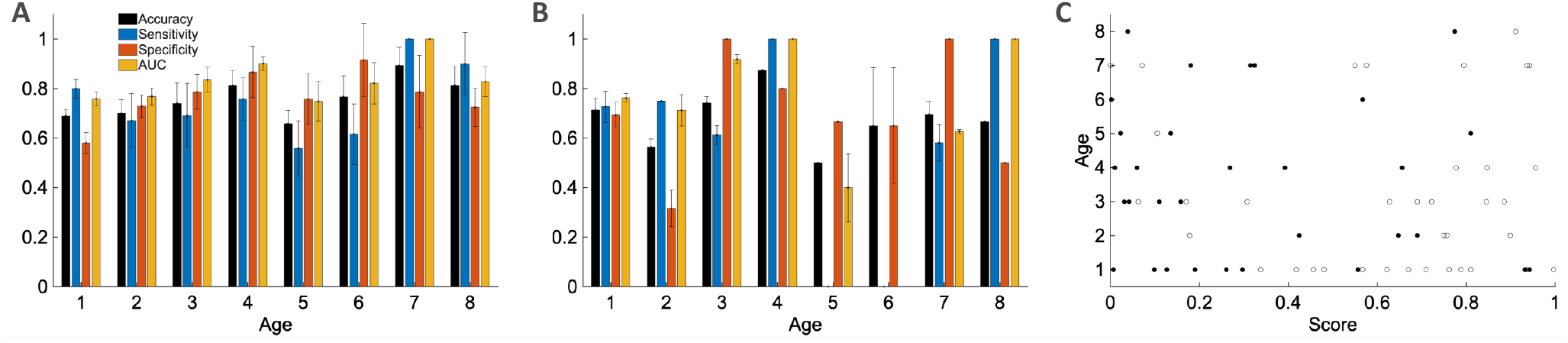
Evaluation of the effect of age. (A) Age-stratified performance metrics in the discovery dataset (accuracy, sensitivity, specificity, and AUC). (B) Age-stratified performance metrics in the validation dataset. (C) Relationship between prediction score and age. White dots indicate KD and black dots indicate non-KD in the validation dataset.

We also examined the effect of body temperature because fever can influence heart-rate and other ECG features [17,18] (Figure 5). In the validation dataset, among patients with body temperature below 38.5°C, the model achieved an accuracy of 0.652 ± 0.018, sensitivity of 0.620 ± 0.024, specificity of 0.690 ± 0.029, and AUC of 0.685 ± 0.019. In the group with body temperature of 38.5°C or higher, the model achieved an accuracy of 0.859 ± 0.044, sensitivity of 0.865 ± 0.075, specificity of 0.850 ± 0.032, and AUC of 0.919 ± 0.024. In the scatter plot of prediction score versus body temperature in the validation dataset, no significant correlation was observed between body temperature and prediction score (r = 0.10, p = 0.41) (Figure 5C).

**Figure 5.**
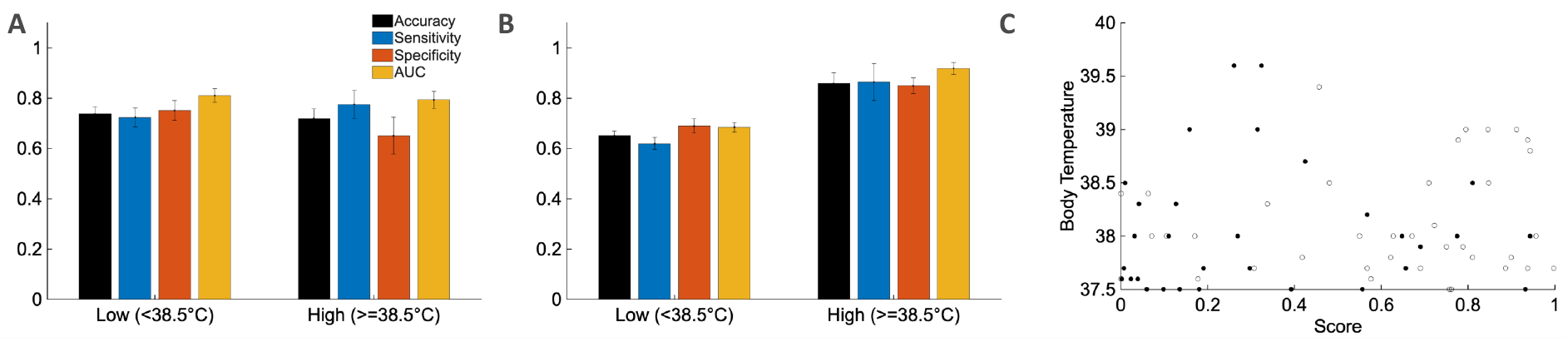
Evaluation of the effect of fever severity. (A) Performance metrics in the discovery dataset for patients with body temperature below 38.5°C and those with body temperature of 38.5°C or higher (accuracy, sensitivity, specificity, and AUC). (B) Performance metrics in the validation dataset for patients with body temperature below 38.5°C and those with body temperature of 38.5°C or higher. (C) Relationship between prediction score and body temperature. White dots indicate KD and black dots indicate non-KD in the validation dataset.

## Discussion

### Principal findings

In this study, we found that a logistic regression model using 11 features extracted from 12-lead ECGs could distinguish pediatric patients with complete KD from pediatric patients with fever. The model achieved an AUC of 0.737 in the validation dataset. In particular, the model showed an AUC of 0.919 for patients with body temperature of 38.5°C or higher, although this subgroup was small and the result should be interpreted cautiously. These results suggest that ECG may serve as an additional source of information in the clinical assessment of KD. Although this level of performance is not sufficient to replace definitive diagnosis, it is clinically meaningful that moderate discrimination was achieved using a rapid, noninvasive test alone. The amplitude of the P, Q, and S waves and the RR interval were also identified as potentially important predictive features.

### ECG features of KD

Recent studies have reported associations between ECG features and KD [5–8]. In our study, S-wave amplitude and the RR interval were the most consistently selected features in top-performing models, followed by P- and Q-wave amplitudes. T-wave amplitude was also selected in a moderate proportion of top-performing feature subsets, whereas R-wave amplitude and most timing features were selected less frequently. The contribution of the RR interval suggests that heart-rate-related autonomic changes during acute inflammation may contribute to discrimination. The QT interval showed a relatively large coefficient in the logistic regression model, possibly reflecting its correlation with the RR interval, but it was less frequently selected in the exhaustive feature-subset analysis. Because the correlation between RR interval and QT interval was about 0.59 (not shown), QT interval may have contributed to the prediction. The repeated selection of P-, Q-, and S-wave amplitudes suggests that waveform morphology in the precordial leads may also contribute. Rather than examining each previously reported index separately, the present study is notable for simultaneously handling multiple amplitude and timing features to extract an electrophysiological pattern associated with KD. The repeated inclusion of P-, Q-, and, S-wave amplitude, and RR interval in top-performing models suggests that the observed signal was multivariable rather than dependent on a single repolarization marker.

### Potential influence of age and fever

Age is a major potential confounder in pediatric ECG analysis because normal intervals, amplitudes, and precordial waveform patterns change substantially across childhood [19–21]. In the present study, we attempted to mitigate this influence by balancing the age distributions of the KD and non-KD groups during model development. In addition, the lack of a significant linear correlation between prediction score and age in the validation dataset suggests that the model output was not driven solely by age-related differences. Importantly, age-stratified analyses still showed discriminative performance across several age groups. This pattern argues against a trivial explanation based solely on differences in age distribution between KD and non-KD groups. Rather, the model appears to capture KD-related ECG information superimposed on normal developmental variation.

Fever is another plausible source of heterogeneity because body temperature affects heart rate and can influence ECG parameters [17,18]. In our fever-stratified analysis, discrimination was preserved in both fever groups, while the group with body temperature ≥38.5°C showed better overall performance. One possible explanation is that greater inflammatory burden accentuates the electrophysiological differences between complete KD and other febrile illnesses, thereby making the ECG signal more separable. However, this higher performance was not accompanied by a trade-off between sensitivity and specificity; both metrics changed in the same direction across fever groups. This pattern argues against a simple prediction bias in which fever severity would merely push the model toward overcalling either KD or non-KD.

### Limitations

This study has several limitations. First, it was a single-center retrospective study, and the sample size remained limited. Second, the training data were restricted to complete KD, and generalizability to incomplete KD has not been sufficiently validated. This limitation is clinically important because incomplete KD remains one of the most difficult diagnostic settings and may lie on a spectrum with complete KD [22]. Third, clinical conditions at the time of ECG acquisition, including day of illness, timing relative to treatment, degree of fever, sedation, and hemodynamic status, could not be fully adjusted for. Nevertheless, an important strength of this study is that the model was developed using definitively diagnosed complete KD and then evaluated in a temporally separate cohort. This design provides an initial assessment of temporal generalizability and supports the potential utility of the model as an adjunctive diagnostic tool.

## Data Availability

The data supporting the findings of this study are not publicly available because they include potentially identifiable clinical and electrocardiographic data from pediatric patients. Data are available upon reasonable request and approval of institutional ethics committee. Analysis codes used in this study will be available.

https://github.com/nakano-lab

## Future directions

Future work should include external validation using multicenter datasets and extension toward severity prediction incorporating clinical outcomes such as coronary artery lesions, intravenous immunoglobulin resistance, and recurrence. Our study used hand-engineered features and logistic regression, an approach that offers the advantage of interpretability. On the other hand, it cannot fully represent local waveform shape or temporal dependencies across multiple leads. With a larger sample size and external validation, it would be worthwhile to investigate deep-learning models that directly use raw ECG waveforms as input. In addition, integration of AI model outputs with blood test findings, day of illness, echocardiographic findings, and existing risk scores may improve both diagnostic performance and clinical implementation.

## Conclusions

Combining features extracted from 12-lead ECGs with machine learning enabled discrimination between complete KD and other pediatric patients with fever. ECG is noninvasive and widely available, and with external validation and multimodal integration, this approach may evolve into an early adjunctive diagnostic tool for KD.

## Acknowledgments

This study was supported by JSPS KAKENHI Grant Number JP26K00592, Suzuken Memorial Foundation, Research Foundation for the Electrotechnology of Chubu, and Aichi Health Promotion Foundation. We also acknowledge Dr. Tadayoshi Hata from Health Care Center, Meiekimae Clinic, Seikokai Medical Corporation, Nagoya, Japan.

## Data Availability

The data supporting the findings of this study are not publicly available because they include potentially identifiable clinical and electrocardiographic data from pediatric patients. Data are available upon reasonable request and approval by the institutional ethics committee. Analysis codes used in this study are available in Github.

